# Social Media Surveillance for Perceived Therapeutic Effects of Cannabidiol (CBD) Products

**DOI:** 10.1101/19004929

**Authors:** Tung Tran, Ramakanth Kavuluru

## Abstract

**Background:** CBD products have risen in popularity given CBD’s therapeutic potential and lack of legal oversight, despite lacking conclusive scientific evidence for widespread over-the-counter usage for many of its perceived benefits. While medical evidence is being generated, social media surveillance offers a fast and inexpensive alternative to traditional surveys in ascertaining perceived therapeutic purposes and modes of consumption for CBD products.

**Methods:** We collected all comments from the CBD subreddit posted between January 1 and April 30, 2019 as well as comments submitted to the FDA regarding regulation of cannabis-derived products and analyzed them using a rule-based language processing method. A relative ranking of popular therapeutic uses and product groups for CBD is obtained based on frequency of pattern matches including precise queries that entail identifying mentions of the condition, a CBD product, and some “trigger” phrase indicating therapeutic use. We validated the social media-based findings using a similar analysis on comments to the U.S. Food and Drug Administration’s (FDA) 2019 request-for-comments on cannabis-derived products.

**Results:** CBD is mostly discussed as a remedy for anxiety disorders and pain and this is consistent across both comment sources. Of comments posted to the CBD subreddit during the monitored time span, 6.19% mentioned anxiety at least once with at least 6.02% of these comments specifically mentioning CBD as a treatment for anxiety (i.e., 0.37% of total comments). The most popular CBD product group is oil and tinctures.

**Conclusion:** Social media surveillance of CBD usage has the potential to surface new therapeutic use-cases as they are posted. Contemporary social media data indicate, for example, that stress and nausea are frequently mentioned as therapeutic use cases for CBD without corresponding evidence, that affirms or denies, in the research literature. However, the abundance of anecdotal claims warrants serious scientific exploration moving forward. Meanwhile, as FDA ponders regulation, our effort demonstrates that social data offers a convenient affordance to surveil for CBD usage patterns in a way that is fast and inexpensive and can inform conventional electronic surveys.

## Introduction

Cannabidiol (CBD) is a well-known cannabinoid compound derived from certain strains of *Cannabis Sativa*. It has been shown to have high potential for therapeutic efficacy and low potential for abuse and dependency in humans (World Health Organization 2017); however, evidence is not substantial enough to warrant wide spread over-the-counter availability of CBD products for perceived therapeutic effects (Cogan 2019, Cohen and Sharfstein 2019). As of now, the U.S. Food and Drug Administration (FDA) has approved a CBD based drug only for epilepsy in June 2018 (U.S. Food and Drug Administration 2018). Nevertheless, public interest in CBD has skyrocketed in recent years. The 2018 United States Farm Bill, which became law on December 20, 2018, included a provision that removed hemp from Schedule 1 controlled substances. As a result, CBD based products, derived primarily from hemp plant extracts, are now ubiquitous in the U.S. marketplace for over-the-counter purchase (backed by major retailers including Walgreens, Kroger, and CVS) and remain relatively unregulated (Reily 2019). Even if there is no long-term harm, aggressive marketing of CBD products (oils, edibles, topicals, and vapes) by the cannabis industry could lead to significant cost burden to consumers as they may regularly buy products for claimed/perceived health benefits without conclusive clinical evidence. Additionally, because of aggressive CBD marketing, consumers may forgo more evidence-based treatments in favor of CBD products.

As more medical evidence is being generated and consolidated, it is critical to keep track of therapeutic purposes that consumers indicate on social media to inform policy and prevention initiatives. In this first of its kind study for CBD, we analyze perceived or expected therapeutic uses for CBD based on social media discussions. Our precise rule-based method is used to ascertain the relative ranking of different therapeutic reasons for the use of CBD according to public perception. We apply a similar method to identify popular ways in which CBD is consumed. Given suitable anonymity properties, we chose the CBD subreddit on the Reddit social platform for this effort. We additionally validate our method on a collection of public comments submitted as part of a recent FDA request for comments (RFC Docket ID: FDA-2019-N-1482) regarding regulation of cannabis-derived products. While traditional surveys and literature reviews are powerful tools for assessing potential uses for CBD, we argue that automated surveillance of social media platforms offer a fast and inexpensive alternative that could inform and complement more traditional sources of surveillance. Specifically, it is possible to continually perform this kind of analysis in a live or streaming fashion. Furthermore, this type of automatic analysis on CBD usage and effects may facilitate the construction and deployment of traditional surveys by helping to populate survey choices.

## Background

Althouse et al. (2015) first introduced the concept of *novel data streams* (NDS) in the context of public health surveillance and defined NDS as data streams with contents that are generated by users (or patients) themselves, such as social media and web search data, and excludes traditional data sources such as electronic health record, disease registries, and vital statistics. Paul et al. (2016) compared information reported in online forums (i.e., NDS data) against large epidemiology survey data and found significant correlations between the two sources; their results suggest that while online forums are not reliable for predicting long-term trends, they are a reliable source for estimating temporal demographic associations and trends in drug use. A recent study to assess the quality of solicited advice for treatment of chronic pain (including treatment using cannabis products) in Reddit comments found that at least 33.9% had at least some support in the research literature (K. L. Costello 2019), suggesting that health-related discussions on Reddit are, to a reasonable extent, substantial and nuanced. Indeed, several studies have analyzed Reddit communities (subreddits) as a means of exploring temporal trends and deriving insights related to use of cannabis products (Sowles, et al. 2017, Meacham, Paul and Ramo 2018, Costello, Martin III and Edwards Brinegar 2017). Costello et al. (2017) specifically found that drug-related subreddits often contain banter (i.e., off-topic discussions) and argues that banter is an essential part of discussions as they foster disclosure and information-seeking behavior. Enghoff and Aldridge (2019) has since argued that unsolicited online data, including Reddit comment data, is increasing valuable in the digital age for the purpose of informing drug policy. To our knowledge, no studies have explored NDS data to determine perceived remedial effects and usage patterns for CBD.

## Materials and Methods

We perform our analysis on two collection of comments from the following sources.

1. A collection of all comments posted to the CBD subreddit between January 1, 2019 and April 30, 2019 totaling to 64,099 individual comments. To keep our methodology simple, we only analyzed comments (as opposed to “posts”), which were treated as independent units of discourse irrespective of the complex threading structure. We sourced Reddit comment data from the *pushshift*.*io* platform^2^ which comprehensively archives Reddit comments on a monthly basis (while trailing behind the “live” data by several months). We filtered for comments specifically posted to the CBD reddit within the target date range, which corresponds to the four-month period roughly between when the 2018 Farm Bill became law and the date of the latest comments available on *pushshift*.*io* at the time of writing.
2. A collection of 3832 machine-processable comments^1^ submitted to FDA’s RFC on cannabis derived products between April 3, 2019 and July 19, 2019. There were 4272 comments in total^1^ by the time the comment period closed on July 19, 2019, and of them 3832 were machine processable comments as ASCII text (mostly from consumers) while the rest included PDF attachments (some paper scans) that were not readily processible, mostly including formal opinions from organizations that are in the stakeholder group for CBD products. We exclusively focused on typed-in comments that were from consumers.

These platforms were chosen for our analyses because of the tendency for comments to be verbose and focused with respect to the topic of cannabis-derived products.

First, we use the MetaMap concept identification and normalization tool (U.S. National Library of Medicine 2019) to identify frequently mentioned psychological and physiological conditions at the concept level. MetaMap is a knowledge-based tool used to identify mentions of medical concepts (e.g., drugs, diseases, and symptoms) in free text and link them to a concept-unique IDs. We specifically considered concepts that are either of semantic type *Mental and Behavioral Dysfunction (MOBD)* or *Disease or Disorder (DSYN)*. Based on the concepts found, we manually curated an exhaustive list of *target conditions*; each target condition is associated with a dictionary of terms (i.e., ways in which a condition is expressed) through manual review of disease and disorder concepts frequently identified by MetaMap. For example, for seizure disorders, we look for mentions of terms such as “epilepsy”, “epileptic”, and “seizure/s”.

As the CBD subreddit may contain “off-topic” discussion of ailments not directly related to CBD, we also experimented with increasingly precise queries, based on regular expression patterns to match comments that specifically mention the target condition as well as CBD and some therapeutic phrase. The three type of queries corresponding to different levels of restrictiveness are as follows. For each comment, we

1. search for mentions of the target condition,
2. search for mentions of the target condition and CBD within the same sentence,
3. search for mentions of the target condition, CBD, and some therapeutic phrase indicating treatment where each mention is separated by at most 36 characters (half the observed average sentence length). We match on several variations of the pattern to allow matching on all possible orders, allow for synonyms of CBD including cannabidiol and hemp oil, and allow for queries based on different therapeutic phrases.

Here, therapeutic phrases come from a hand-crafted lexicon of 87 terms including “treats”, “cures”, “helps”, “reduces”, “alleviates”, “relieves”, “eliminates”, “kills”, “stops”, “eases”, “aids”, “soothes”, “inhibits”, “improves”, “destroys”, “reverses”, “suppresses”, “lowers”, “regulates”, “prevents”, “manages”, “fixes”, “better” and their variants including conjugated forms. The lexicon was developed by manually examining samples of comments that matched queries (1) and (2). From the initial list of phrases, we added several more after consulting an online thesaurus for likely synonyms. The queries are performed in a case insensitive fashion and matches containing negation terms (including “never”, “not”, and related contractions) are disregarded. We emphasize that *frequency* or *count* is defined as the number of *unique* comments containing the term or matching a pattern-based query; that is, a term or pattern match is only counted at most once per comment even if it occurs multiple times in some comments. Comments mentioning multiple conditions will increment the match count for each condition separately. For example, a comment containing the statement “CBD helps my anxiety and pain” would increment the match count for both *anxiety* and *pain* at all levels of restrictiveness. By manually examining 100 randomly sampled matches at the strictest (third) query type, we estimate our method to be 96% accurate (more precisely, 96% positive predictive value rate) with a 95% confidence interval of [90.16%, 98.43%]. However precise, we note that the frequency of pattern matches is an underestimate of the true frequency.

To explore the modes of consumption for CBD products, we curated a list of popular CBD products based on a manual review of popular CBD e-commerce websites. We group them into five broad product groups including Oils/Tinctures, Vapes, Edibles, Pills/Capsules, and Topicals. Less obvious terms were obtained by querying similar word vectors as induced by the distributional semantics software Word2Vec (Google Code Archive 2013) on the comment data; for example, by querying for words similar to “vape”, “vaping”, we obtained additional terms such as “dab” and “wax” which are associated with a less well-known mode of inhalation. We note that mentions of vaping, for example, are not all CBD-related; some posts are simply discussing the default practice of vaping *nicotine liquid*. As such, we additionally perform more precise queries to match for mentions of CBD products that explicitly contain as a prefix “CBD”, “cannabidiol”, or “hemp” such as “CBD tea” or “hemp lotion.”

## Results and Discussion

We report our analysis of top ten mentioned conditions in Table 1, sorted by matching at the strictest level on Reddit comments, where the second column is a list of terms for the target condition along with their individual frequencies (in parentheses). The next six columns are frequencies for matches of queries at varying levels of restrictiveness, based on pattern rules, with an example at the last column. We present a plot of the proportions for each condition along with their 95% confidence intervals in Figure 1. Based on these results, anxiety disorders and pain are the two conditions dominating much of the discussion surrounding CBD, both in terms of general discussion and as a perceived therapeutic treatment. This is consistent for both comments posted to the CBD subreddit and comments submitted to the FDA’s 2019 RFC on cannabis-derived products.

**Table 1.** Most frequently mentioned conditions in the CBD subreddit. Superscript ^(A)^ marks conditions discussed in a review of studies regarding CBD therapeutic efficacy (White 2019). Superscript ^(B)^ marks conditions discussed in a recent survey study of CBD users (Corroon and Phillips 2018).

| Conditions | Term Count<br>(Reddit Only) | Reddit Comments |  |  | FDA RFC Responses |  |  | Example Comments with Mentions including Target Condition, CBD, and Treatment Phrase |
| --- | --- | --- | --- | --- | --- | --- | --- | --- |
|  |  | Mentions Count | Mentions with CBD Count | Mentions with CBD and Treatment Phrase Count | Mentions Count | Mentions with CBD Count | Mentions with CBD and Treatment Phrase Count |  |
| **Anxiety Disorders** <sup>(A)(B)</sup> | anxiety (3596)<br>anxious (376)<br>panic attacks (323)<br>social anxiety (138)<br>anxiety attack (60)<br>general anxiety (41)<br>GAD (39)<br>panic disorder (33)<br>anxiety/depression (32)<br>anxiety symptoms (15)<br>generalized anxiety disorder (12)<br>anxiousness (9)<br>general anxiety disorder (6) | 3968 | 872 | 239 | 1033 | 326 | 159 | I use **CBD** for **anxiety** and personally feel like I've been getting **better** results with this vape setup than I ever did taking a full spectrum tincture. |
| **Pain** <sup>(A)(B)</sup> | pain (2052)<br>chronic pain (189)<br>back pain (127)<br>nerve pain (56)<br>painful (38)<br>breakthrough pain (29)<br>ache (19) | 2068 | 419 | 115 | 1823 | 770 | 295 | I find that for me, vaping **CBD** in the morning and evening **helps** my **back pain** and sciatica. |
| **Depressive Disorders** <sup>(B)</sup> | depression (592)<br>depressed (63)<br>anxiety/depression (32)<br>miserable (22)<br>depressing (8)<br>misery (5) | 667 | 110 | 36 | 301 | 59 | 30 | Thank you for the review, I just bought some **CBD** flower and I'm hoping that it will **help** with my **depression** and get me back to be productive. |
| **Inflammation** <sup>(B)</sup> | inflammation (307)<br>inflammatory reaction (2) | 309 | 87 | 31 | 217 | 65 | 28 | The **CBD** **reduces** my **inflammation** so having my inflammation reduced for a period of time sense to make it so my pain doesn't spike as high long-term. |
| **Stress** | stress (347)<br>stressful (66)<br>stressed (60)<br>stressing (11)<br>stresses (8) | 465 | 58 | 16 | 187 | 32 | 14 | Because autism seems so complex and I only saw **CBD** being talked about for **stress** I thought it just **lowered** stress but it really reduces the feelings of overstimulation. |
| **Headache** <sup>(A)(B)</sup> | headaches (286)<br>migraines (222) | 431 | 78 | 12 | 204 | 65 | 27 | **CBD** has been the best thing yet in **reducing** nicotine cravings and **headaches** I have found. |
| **Sleep Disorders** <sup>(A)(B)</sup> | insomnia (206)<br>sleep issues (20)<br>sleep disorders (13)<br>sleep problems (12)<br>sleeping problems (10)<br>problems sleeping (3)<br>sleepless (3)<br>sleeplessness (2) | 265 | 52 | 12 | 233 | 40 | 21 | **CBD** **took care of** my chronic **insomnia** issues, and I no longer need Zolpimist for sleep. |
| **Seizure Disorders** <sup>(A)(B)</sup> | seizures (199)<br>epilepsy (109)<br>epileptic (20) | 277 | 63 | 9 | 236 | 79 | 37 | Also, anecdotally, I'm told by a doctor that gives CBD to epileptic children that full-spectrum is more effective than **CBD** isolate (for **eliminating seizures**, at least). |
| **Cancer** <sup>(B)</sup> | cancer (233)<br>cancerous (12) | 244 | 41 | 9 | 215 | 39 | 9 | I know when my cousin was using **CBD** oil to **treat** his **cancer** the doctors and nurses were almost rude to him. |
| **Nausea** | nausea (128)<br>nauseous (21)<br>nauseated (7)<br>nauseating (4) | 157 | 24 | 9 | 73 | 6 | 2 | My aunt was diagnosed with CHS seven years ago and aside from hot showers the only thing that actually **relieves** her **nausea** is **CBD**. |

**Figure 1.**
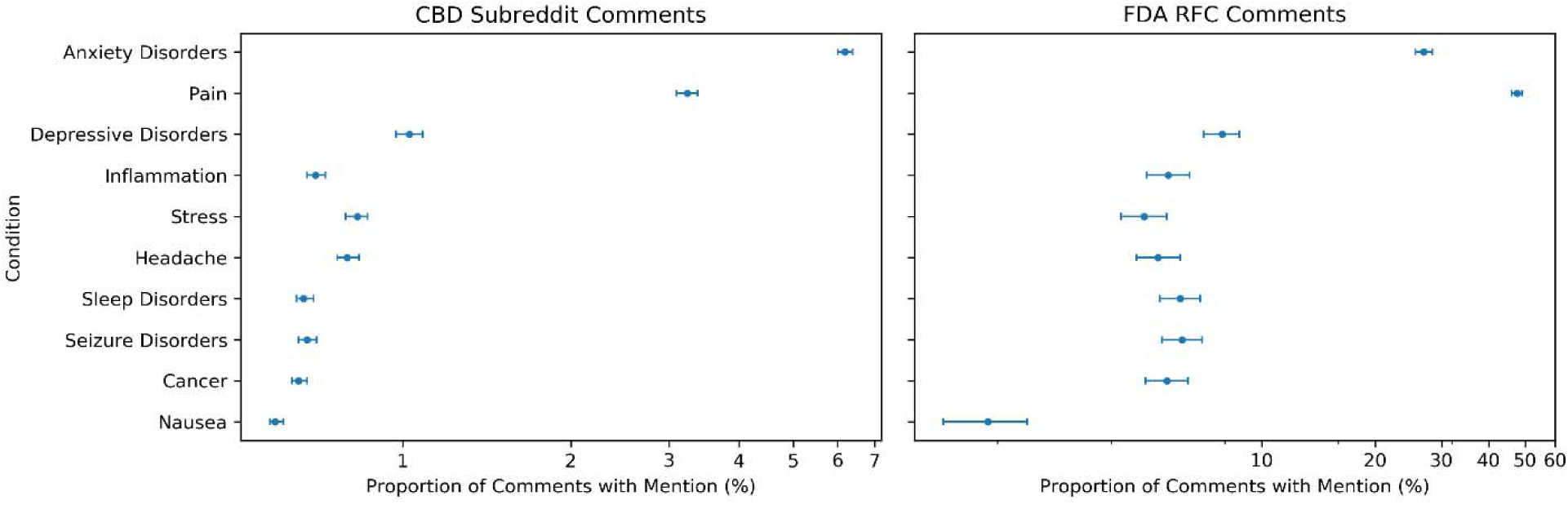
95% confidence intervals around the proportion of total comments mentioning each condition.

Despite having an order of magnitude more Reddit comments than comments to the FDA, we note that there is not a dramatic difference in the number of term/pattern matches between the two platforms. This may indicate that comments to the FDA, as expected, are highly focused on the perceived therapeutic effects of CBD products while Reddit comments are more likely to include off-topic discussions. Given the match counts/frequencies are comment-unique (and underestimates the true frequency), it is possible to assess, for example that 6.19% (3968 out of 64099) of comments posted to the CBD subreddit mention anxiety, and that at least 6.02% (239 out of 3968) of these comments explicitly discuss CBD as a potential or perceived remedy for anxiety (0.37% of total comments); for pain, these percentages are 3.22% and 5.56% respectively (0.17% of total comments). On the other hand, 27.00% of comments to the FDA RFC mention some form of anxiety disorder, and at least 15.39% of these comments explicitly mention CBD as a potential therapeutic solution (4.16% of total comments); for pain, these percentages are much more prominent at 47.57% and 16.18% respectively (7.60% of total comments).

From Table 1, it can be surmised that Reddit comments tend to focus more on mental conditions while FDA comments tend to focus more on physiological conditions. To assess this, we performed the original experiment on groups of conditions (from Table 1) organized based on whether they are in the mental (anxiety, depression, stress) or physiological (pain, inflammation, headache, sleep disorder, seizure disorders, nausea, and cancer) category. Reddit comments had mentions of mental conditions in at least 4373 comments (6.82% with a 95% confidence interval of [6.63%, 7.02%]) and physiological conditions in at least 3262 comments (5.09% with a 95% confidence interval of [4.92%, 5.26%]). On the other hand, FDA comments had mentions of mental conditions in at least 1191 comments (31.02% with a 95% confidence interval of [29.63%, 32.56%]) and physiological conditions in at least 2260 comments (58.98% with a 95% confidence interval of [57.41%, 60.52%]). The reported distributions seem to affirm that FDA comments tend to focus heavily on physiological conditions (compared with mental conditions), while Reddit comments tend to focus only slightly more on mental conditions (compared with physiological conditions). The difference in anonymity provided by these platforms and the greater stigma associated with disclosure of mental health conditions may be one explanation for the discrepancy; while FDA comments may be submitted anonymous, most commenters (82%) chose to submit comments with a partial or full name.

We use superscript (A) in Table 1 to indicate conditions covered in a recent review of human studies assessing the potential of CBD (White 2019) and superscript (B) in Table 1 to indicate conditions covered in a recent survey study of CBD users (Corroon and Phillips 2018). Among conditions frequently discussed on social media that are **not** discussed in research literature are stress and nausea. Less frequently discussed conditions (not shown in Table 1) for which there is little or no research evidence (as observed from PubMed searches) include ADHD and autism, with users making comments such as, “I will say from my personal experience that hemp flower and oil have really helped my ADHD.”

We similarly report popular modes of consumption in Table 2. We found that CBD oil and tinctures are most popular, either as food additives or directly administered sublingually, with vaping being the second most popular mode of consumption. Approximately 13% of the Reddit comments mention oil or tinctures, and 25% of these are explicitly mentioned as a CBD product (3.25% of total comments). In FDA comments, oil and tinctures are likewise most popular at 1442 mentions; however, vapes were mentioned the least number of times at a frequency of 87. Other modes of consumption, including topicals, pills/capsules, and edibles, are mentioned in at similar levels of frequency at 319, 295, and 254 matches respectively.

**Table 2.** Popular modes of consumption based on product group mention frequency in the CBD subreddit.

| Product Groups | Terms | Mentions Count | Mentions with CBD Prefix | Mentions with CBD Prefix Count |
| --- | --- | --- | --- | --- |
| **Oil/Tinctures** | oil (6461)<br>tincture (2687)<br>drops (1143)<br>droplets (12) | 8609 | CBD oil (1701)<br>hemp oil (374)<br>CBD tincture (193)<br>CBD drops (29) | 2215 |
| **Vapes** | vape (3684)<br>juice (955)<br>vape juice (394)<br>dab (360)<br>wax (340)<br>vape pen (278)<br>pods (190)<br>ejuce (180)<br>vape cartridge (124)<br>eliquid (101)<br>vape liquid (40)<br>vape tank (19) | 4775 | CBD vape (277)<br>CBD vape juice (88)<br>CBD juice (41)<br>CBD ejuce (26)<br>CBD dabs (24)<br>CBD pods (20)<br>CBD eliquid (19)<br>CBD vape pen (19)<br>CBD wax (13)<br>CBD vape cartridges (10)<br>CBD vape liquid (8)<br>hemp pods (6)<br>hemp vape (4) | 419 |
| **Edibles** | edibles (748)<br>gummies (688)<br>coffee (337)<br>candy (249)<br>tea (245)<br>chocolate (122)<br>drinks (96)<br>honey (71)<br>cookies (70)<br>brownies (38)<br>syrup (34)<br>beverage (30)<br>snacks (30)<br>smoothies (16)<br>chews (11) | 2429 | CBD gummies (81)<br>CBD edibles (56)<br>CBD coffee (20)<br>CBD tea (14)<br>hemp tea (9)<br>hemp edibles (8)<br>CBD drinks (4)<br>CBD honey (4)<br>CBD candy (4)<br>hemp honey (3)<br>hemp gummies (2)<br>CBD snacks (2)<br>CBD beverages (2) | 209 |
| **Pills/Capsules** | capsules (726)<br>pills (445)<br>caps (127)<br>tablets (93)<br>softgels (15) | 1270 | CBD capsules (92)<br>CBD pills (15)<br>CBD caps (11) | 128 |
| **Topicals** | cream (228)<br>salve (166)<br>balm (163)<br>topicals (115)<br>lotion (102)<br>patches (76)<br>gel (46)<br>ointment (13)<br>topical cream (12)<br>body butter (12) | 787 | CBD balm (22)<br>CBD cream (17)<br>CBD salve (12) | 96 |

### Limitations and Ethics

Given the precise nature of our methodology (and rule-based methods in general), our results in fact lead to underestimates of proportions of Reddit posters perceiving therapeutic potential. However, we contend that our method is sufficient as the goal is to obtain a relative ranking among popularly discussed conditions and not necessarily to obtain absolute estimates of CBD users using it for a particular therapeutic reason. As a limitation, our analyses and estimates do not necessarily reflect the perception and usage patterns of the general population of CBD users, who may or may not comment in these specific forums. Such an estimate can only be obtained by a conventional survey targeting a truly random sample of consumers and not just those who post on Reddit or submit comments to the FDA. However, our effort has the potential to surface new therapeutic use-cases (as they are posted), which can then be included as options in more traditional surveys that can be timed at regular intervals as per resource constraints.

The ethics of using social media data in research is still a contentious topic. Given their private yet public nature, some researchers argue that social media data should be treated as public data that can used without ethical considerations while others argue that it should be regarded as research with human participants and thus treated with the appropriate ethical stipulations (Enghoff and Aldridge 2019). Herein, we address ethical concerns related to our methodology. First, we note that, unlike many social media platforms, Reddit users typically use pseudonyms or handles not related to their real identity. As of the time of this writing, only a username and password and no other identifying information are required for registration to the platform. Second, we note that data used in our research is *openly and publicly available* in that no form of registration or admittance is required to access them. In example comments shown in Table 1, we have changed minor details to eliminate any potential for identification. Lastly, besides providing anonymized examples of comments for context, we report only aggregate data. Thus we believe that our study conforms to common guidelines for ethical research using online social media data (Markham and Buchanan 2012, Gonzalez-Hernandez 2019).

We note that online groups and communities may feel ownership over the content they produce. While effort has been taken to ensure anonymity, the analyses in this study were performed without input from individual users, content moderators, or community leaders. While permission cannot be obtained post-factum, in future studies of a similar nature, we recommend reaching out to community leaders and content owners with offers of research collaboration early on. In doing so, researchers exercise best practices for social media data research.

## Conclusion and Future Work

CBD’s fast-growing popularity fueled by the current relatively unregulated landscape warrants a serious and continuous exploration of perceived therapeutic claims by consumers. We took a first step in that direction in this effort by mining CBD related social media chatter. Specifically, we analyzed social media comments to the Reddit platform and consumer comments to the 2019 FDA RFC regarding cannabis product regulation to attain a relative ranking of perceived therapeutic uses for CBD. We analyzed these comments at varying levels of granularity with respect to a target condition, including assessing whether the comment simply mentioned the target condition or if there was an explicit mention of therapeutic effect related to CBD usage. Additionally, using a similar method, we obtain a relative ranking of popular CBD products as measured by discussion frequency. To our knowledge, this is the first effort to mine CBD related online content. In future efforts, we will explore the use of smarter and less mechanical methods, such as machine learning classifiers, to recognize mentions of conditions and products. We believe such an approach will be less likely to underestimate the true frequencies. Moreover, we will explore the use of fuzzy matching (or spelling correction as a preprocessing step) as a clever way to improve the sensitivity of the proposed method. As FDA ponders regulation, we believe our effort demonstrates that social data offers a convenient affordance to surveil for CBD usage patterns in a way that is fast and inexpensive and can be deployed in a live fashion, offering unique complementary advantages to more traditional surveys.

## Data Availability

Not applicable

## Acknowledgments

This work is supported by the U.S. National Cancer Institute through NIH grant R21CA218231. The content is solely the responsibility of the authors and does not necessarily represent the official views of the NIH.

## Footnotes

1 https://www.regulations.gov/docket?D=FDA-2019-N-1482

2 https://files.pushshift.io/reddit/comments/

